# Few-Shot Learning for Prostate Cancer Detection on MRI: Comparative Analysis with Radiologists’ Performance

**DOI:** 10.1101/2025.01.19.25320503

**Authors:** Yosuke Yamagishi, Yasutaka Baba, Jun Suzuki, Yoshitaka Okada, Kent Kanao, Masafumi Oyama

## Abstract

**Background:** Deep-learning models for prostate cancer detection often require large datasets, which can be challenging to obtain and may lead to domain shift issues in various clinical settings.

**Purpose:** This study aimed to develop a deep-learning model for prostate cancer detection on magnetic resonance images using few-shot learning and compare its performance with radiologists.

**Materials and Methods:** This retrospective study used 99 cases (80 positive, 19 negative) of confirmed prostate cancer, diagnosed through needle biopsy from 2017 to 2022, with 20 cases for training, 5 for validation, and 74 for testing. The 2D transformer model was trained on T2-weighted, diffusion-weighted, and apparent diffusion coefficient map images. Model predictions were compared between the two radiologists using the Matthews correlation coefficient (MCC) and F1 score, and the bootstrap method was used to calculate 95% confidence intervals (CIs).

**Results:** Seventy-four patients (mean age, 71 years ± 8; 60 men) were included in the test set. The model achieved an MCC of 0.297 (95% CI: 0.095–0.474) and F1 score of 0.707 (95% CI: 0.598–0.847). Radiologist 1 had an MCC of 0.276 (95% CI: 0.054–0.484) and an F1 score of 0.741 (95% CI: 0.632–0.832), while Radiologist 2 had an MCC of 0.504 (95% CI: 0.289–0.703) and an F1 score of 0.871 (95% CI: 0.800–0.931). The performance of the model was not significantly different from that of Radiologist 1 (MCC difference: 0.021, 95% CI: −0.270–0.306; F1 score difference: −0.034, 95% CI: −0.153–0.078), but was lower than that of Radiologist 2 (F1 difference: −0.16, 95% CI: −0.287– - 0.061).

**Conclusion:** A deep-learning model trained on only 20 cases achieved a performance comparable to one radiologist in detecting prostate cancer on magnetic resonance images, demonstrating the potential of few-shot learning in addressing domain shift challenges.

**Key Results:**

1. A deep learning model for prostate cancer detection on MRI was developed using only 20 training cases.
2. The model achieved performance comparable to one radiologist (MCC: 0.297 vs 0.276) but lower than another (F1: 0.707 vs 0.871).
3. Few-shot learning demonstrated potential for addressing domain shift challenges in medical imaging AI.

**Summary Statement:** Few-shot learning enables development of prostate cancer detection models on MRI with performance comparable to radiologists, using minimal training data.

## Introduction

The development of deep-learning models for prostate cancer detection has progressed rapidly in recent years. Many researchers have constructed high-performance models using large-scale datasets, paving the way for clinical applications (1–6). In many cases, these models have demonstrated performance comparable to or surpassing that of radiologists (7).

However, in the field of medical image diagnosis, domain shift can occur owing to various factors, such as differences in imaging equipment, patients’ racial backgrounds, and variations in imaging protocols across facilities (8). This domain shift can cause models that perform well in one environment to underperform significantly in another.

Therefore, it is crucial to fine-tune the models at each facility and adapt them to specific environments. However, collecting cases at medical institutions is often difficult and building large-scale datasets is not easily achievable. Therefore, it is extremely important to perform fine-tuning using a small number of cases. Previous studies have investigated the relationship between the model performance and the amount of training data (7). However, research directly comparing the model performance under few-shot learning conditions with the diagnostic accuracy of radiologists is limited.

To address these challenges, we propose an enhanced approach that maximizes the utility of limited magnetic resonance images (MRI) datasets. Our method expands upon established methodologies and previous techniques by adopting a 2D model to leverage the multi-slice nature of MRI data (9), effectively increasing the amount of training data per case. We streamlined the data preparation process using an efficient slice-by-slice labeling system based on the biopsy results.

Despite working with only 20 cases, our goal was to develop a deep-learning model that achieves diagnostic accuracy comparable to that of experienced radiologists. We investigated the effectiveness of fine-tuning techniques using limited data and explored how these models can be realistically implemented in clinical settings.

Herein, we present a method for each medical facility to efficiently optimize models using their own data, thereby improving the diagnostic accuracy and efficiency. This will provide a path for solving the problem of domain shift while reducing the costs associated with large-scale data collection and labeling.

By demonstrating that a model can be developed with minimal data from a facility, we demonstrated that optimal model development is possible for each facility, regardless of the domain shift. Consequently, we expect that any facility can develop a model to enhance diagnostic accuracy and efficiency by collecting a small number of cases and performing simple labeling.

## Materials and Methods

This retrospective study was conducted at a single institution and approved by the Institutional Review Board of our institution. This study adhered to the guidelines outlined in the Checklist for Artificial Intelligence in Medical Imaging (CLAIM) 2024 update (10).

### Dataset

We used multiparametric MRI images comprising T2-weighted images (T2WI), diffusion-weighted images (DWI), and apparent diffusion coefficient (ADC) maps acquired at our hospital between 2017 and 2022. MRI examinations were performed using a Philips Achieva scanner (Philips, Amsterdam, Netherlands) between April 2017 and July 2022.

We randomly selected 100 cases, all of which had undergone MRI-targeted prostate biopsy with a subsequent pathological diagnosis.

One case was excluded because of difficulty in evaluation owing to bleeding from the pre-MRI biopsy. The remaining 99 cases were divided into the training and test datasets. The training dataset was further divided into training and validation subsets. Figure 1 shows a flowchart of patient selection and data distribution.

**Figure 1:**
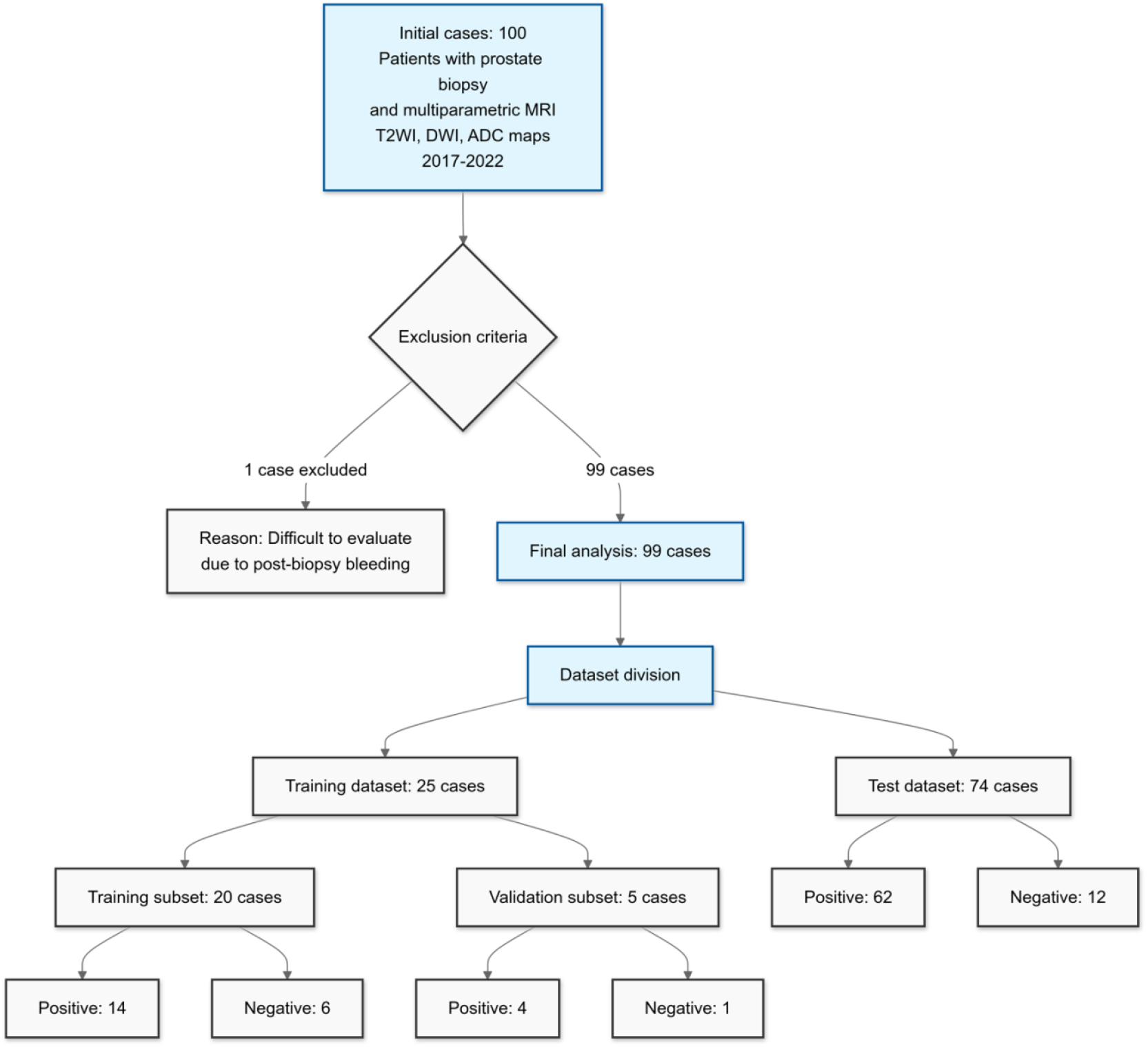
Flowchart of patient inclusion and data distribution in the study of multiparametric MRI data for prostate cancer detection. The flowchart illustrates the selection process from 100 initial cases to 99 analyzed cases, with one patient excluded because of post-biopsy bleeding. This shows the division into training (n = 25) and test (n = 74) datasets, with further subdivision of the training set into training (n = 20) and validation (n = 5) subsets. Positive and negative biopsy results are obtained for each patient subset. MRI, magnetic resonance imaging; T2WI, T2-weighted image; DWI, diffusion-weighted image; ADC, apparent diffusion coefficient.

### Data Preprocessing

The magnetic resonance (MR) image data varied in resolution, ranging from 224 × 224 to 1,008 × 1,008 pixels. To standardize the input, all the images were resized to 224 × 224 pixels using bilinear interpolation. The pixel values were normalized to 256 levels (0-255 for each pixel).

The model input consisted of T2WIs, DWIs, and ADC maps stacked in the channel direction as NumPy arrays (11).

### Model Training

An overview of the proposed approach is shown in Figure 2.

**Figure 2:**
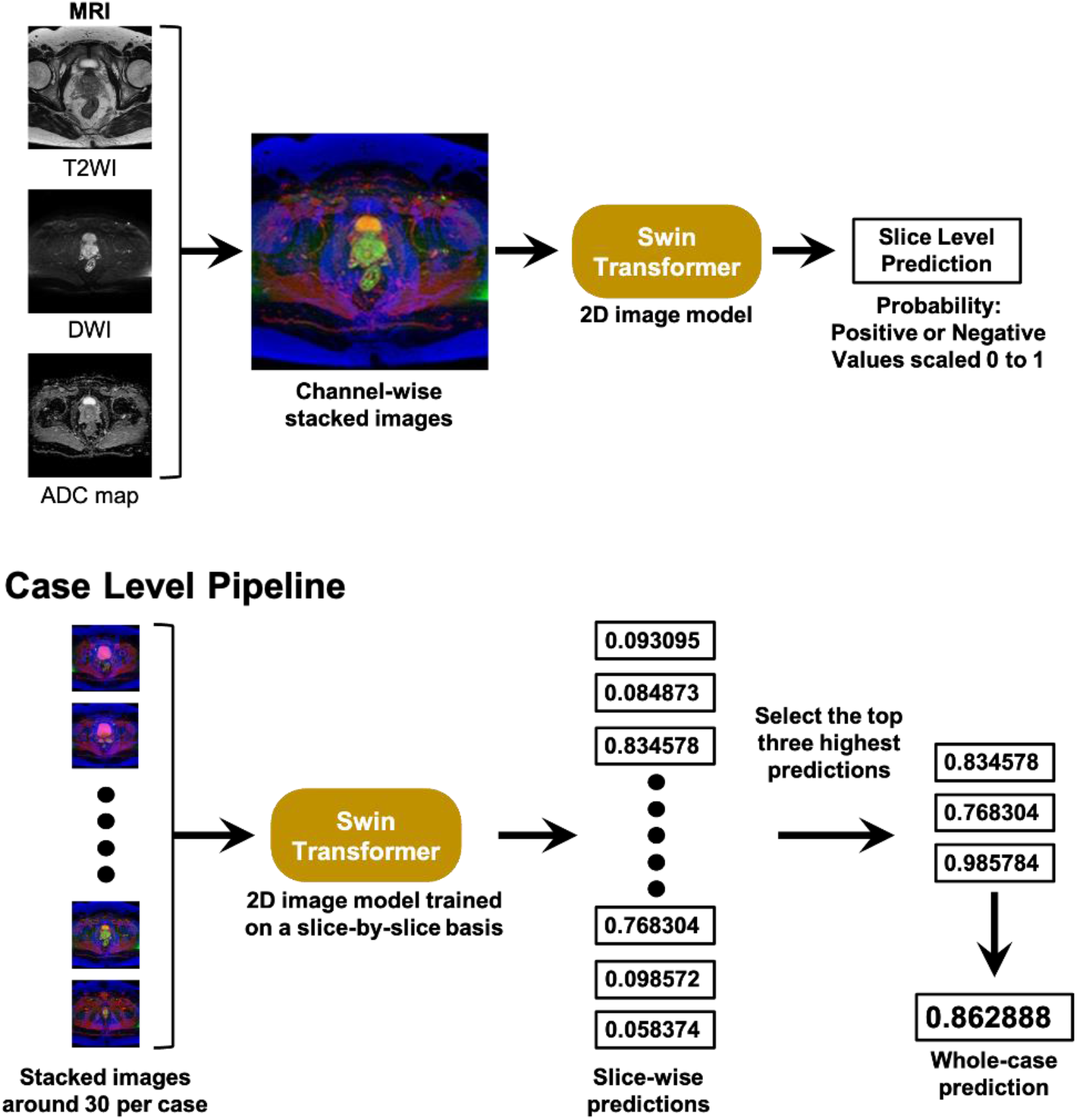
MR image analysis pipelines. This figure presents two interconnected pipelines for magnetic resonance (MR) image analysis: the upper slice-level pipeline for training processes of individual MR image slices through channel-wise stacking, and a Swin Transformer, producing slice-level predictions. The lower case-level prediction aggregates multiple stacked images, processes them through a slice-trained Swin Transformer, and selects the top three predictions to generate a case-level prediction. Both pipelines use the same 2D image models and output probability values from 0 to 1 for positive/negative predictions. MRI, magnetic resonance imaging; T2WI, T2-weighted image; DWI, diffusion-weighted image; ADC, apparent diffusion coefficient.

To train the model with a small number of MR images, we adopted a 2D model instead of 3D models such as 3D convolutional neural network (CNN) models (12), which are frequently used in conventional MR image classification. Using a 2D model, the amount of data input to the model was increased by labeling each slice. The training data were labeled for each slice, depending on the presence or absence of lesions, and 645 2D image data points (96 lesion-positive, 549 lesion-negative) were obtained.

We used the Swin Transformer Small (13) with a patch size of 4 and an image size of 224. The pre-trained weights of the model were initialized using the ImageNet-22k dataset (14) and fine-tuned on the ImageNet-1k dataset (15), which is publicly available in the Timm library (16).

During training, we used a batch size of 8 and trained the model for 10 epochs. The AdamW optimizer (17) was employed, and a warm-up strategy was applied to gradually increase the learning rate to 0.0001 during the first epoch, followed by gradual decay for the remaining 9 epochs. A binary cross-entropy loss function was used.

The Albumentation library (18) was used for data augmentation during the training. The augmentation techniques applied included random resizing and cropping between 85% and 100% of the original scale, random rotation with a maximum angle of 15 °, random horizontal and vertical flipping, random hue and saturation shifts, and cutout with a maximum of 5 randomly placed black patches up to 5% of the image size. After applying these data augmentations, normalization was performed using the mean and standard deviation of the RGB values from the ImageNet dataset, and the normalized images were used as inputs to the model.

The values predicted by the model were obtained for each slice. The three highest predicted values for each case were extracted and their average values were calculated as the predicted values for each case.

The experiments were conducted using Python 3.10 and PyTorch 2.0 (19).

### Statistical Analysis

The performance of the obtained model is evaluated using multiple metrics. The area under the receiver operating characteristic curve (ROC-AUC), precision, recall, and accuracy were calculated. However, to address the challenges of imbalanced datasets, we primarily focused on the Matthews correlation coefficient (MCC) and the F1 score for performance comparisons. All statistical metrics were calculated using the scikit-learn library (20).

The bootstrap method was used to calculate the 95% Credible intervals (CIs) for all metrics. The performance of the model, particularly in terms of F1 score and MCC, was compared with the reading results of two radiologists (hereinafter referred to as Radiologist 1 and Radiologist 2, with 15 and 8 years of experience, respectively) to assess its potential clinical utility using a previously validated method (21).

### Visualization of Model Attention

In addition, gradient-weighted class activation mapping (GradCAM) (22) was used to visualize the importance of pixels contributing to the prediction of a particular class using the gradient information of the model. The gradient of the model output was calculated and weighted for each channel of the feature map. The weighted feature maps for each channel were then summed to produce an importance map for the prediction results.

## Results

### Dataset Characteristic

Table 1 shows that all study participants were Asian, with a mean age of 71.39 years (standard deviation [SD] 7.43) overall, while the training group had a mean age of 73.12 years (SD 7.18) and the test group had a mean age of 70.81 years (SD 7.47). Regarding Gleason scores, 19.2% (19/99) of participants had no cancer. It should be noted that all cases were patients who required biopsy due to suspected prostate cancer, resulting in this high cancer detection rate (80.8%). In the training group, Gleason score 8 (4+4) was most common at 44.0% (11/25), while the test group showed a more balanced distribution across scores from 6 to 10, with Gleason 7 being most frequent (35.1% combined for 3+4 and 4+3).

**Table 1.** Demographic Characteristics of the Study Participants Overall and by Group Allocation.

| Characteristic | Overall (N=99) | Training Group (N=25) | Test Group (N=74) |
| --- | --- | --- | --- |
| Age, years, mean ± SD | 71.39 ± 7.43 | 73.12 ± 7.18 | 70.81 ± 7.47 |
| Race, n (%) |  |  |  |
| Asian | 99 (100%) | 25 (100%) | 74 (100%) |
| Gleason Score |  |  |  |
| No cancer | 19 (19.2%) | 7 (28.0%) | 12 (16.2%) |
| 3 + 3 = 6 | 7 (7.1%) | 0 (0%) | 7 (9.5%) |
| 3 + 4 = 7 | 17 (17.2%) | 3 (12.0%) | 14 (18.9%) |
| 4 + 3 = 7 | 13 (13.1%) | 1 (4.0%) | 12 (16.2%) |
| 4 + 4 = 8 | 22 (22.2%) | 11 (44.0%) | 11 (14.9%) |
| 4 + 5 = 9 | 16 (16.2%) | 3 (12.0%) | 13 (17.6%) |
| 5 + 4 = 9 | 4 (4.0%) | 0 (0%) | 4 (5.4%) |
| 5 + 5 = 10 | 1 (1.0%) | 0 (0%) | 1 (1.4%) |

The actual imaging parameters achieved across all examinations were as follows: slice thickness was consistently maintained at 3.0 mm for all sequence types. The detailed parameters obtained for each sequence are summarized in Table 2, including TR, TE, field of view, and matrix size.

**Table 2:** Distribution of MRI Studies and Imaging Parameters for Training and Test Datasets: Number of Studies by Tesla Strength (Upper) and Mean Imaging Parameters with Ranges (Lower)

|  | Training Case | Test Case |
| --- | --- | --- |
| 3 Tesla studies | 17 | 55 |
| 1.5 Tesla studies | 8 | 19 |
| Parameter |  |  |
| T2WI TR (ms) | 5,612 (3,500–6,550) | 5,672 (4,048–7,604) |
| DWI/ADC map TR (ms) | 6,332 (6,331–7,500) | 6,270 (5,750–7812) |
| T2WI TE (ms) | 96.8 (90.0–100.0) | 97.6 (90.0–100.0) |
| DWI/ADC map TE (ms) | 78.7 (74.7–83.0) | 77.8 (74.4–87.3) |

Note—T2WI, T2-weighted image; TR, repetition time; DWI, diffusion-weighted image; ADC, apparent diffusion coefficient; TE, echo time.

### Model Performance and Comparison with Radiologists

The constructed model for prostate cancer classification demonstrated a significant predictive capability. Its performance metrics were as follows: accuracy, 0.611 (95% CI: 0.500 to 0.723); precision, 0.947 (0.865 to 1.00); recall, 0.567 (0.439 to 0.690); ROC-AUC, 0.730 (0.588 to 0.847); MCC, 0.297 (0.095 to 0.474); and F1 score, 0.707 (0.598 to 0.847).

For comparison, we evaluated the performance of the two radiologists in the same cases. Radiologist 1 achieved an accuracy of 0.641 (0.528–0.750), precision of 0.929 (0.840–1.00), recall of 0.619 (0.492–0.742), MCC of 0.276 (0.054–0.484), and F1 score of 0.741 (0.632–0.832). Radiologist 2 showed a higher performance level, with an accuracy of 0.803 (0.708–0.889), precision of 0.961 (0.898–1.00), recall of 0.797 (0.698–0.889), MCC of 0.504 (0.289–0.703), and F1 score of 0.871 (0.800–0.931).

To compare the performance of the model with that of radiologists, we focused on the differences in the MCC and F1 scores. When compared to Radiologist 1, the model showed a slightly higher MCC (difference of 0.021, 95% CI: −0.270 to 0.306) and a slightly lower F1 score (difference of - 0.034, 95% CI: −0.153 to 0.078), but there were no statistically significant differences. In comparison with Radiologist 2, the MCC of the model was lower, but not significantly (difference of −0.207, 95% CI: −0.480 to 0.060), while its F1 score was significantly lower (difference of −0.16, 95% CI: −0.287 to −0.061).

The results are summarized in Table 3 and Figure 3, which provide a comprehensive comparison of the performance metrics between the constructed model and the two radiologists. The analysis suggests that while the model’s performance was comparable to that of Radiologist 1, there is room for improvement to match the higher performance level demonstrated by Radiologist 2, particularly in terms of overall accuracy and recall.

**Table 3:**
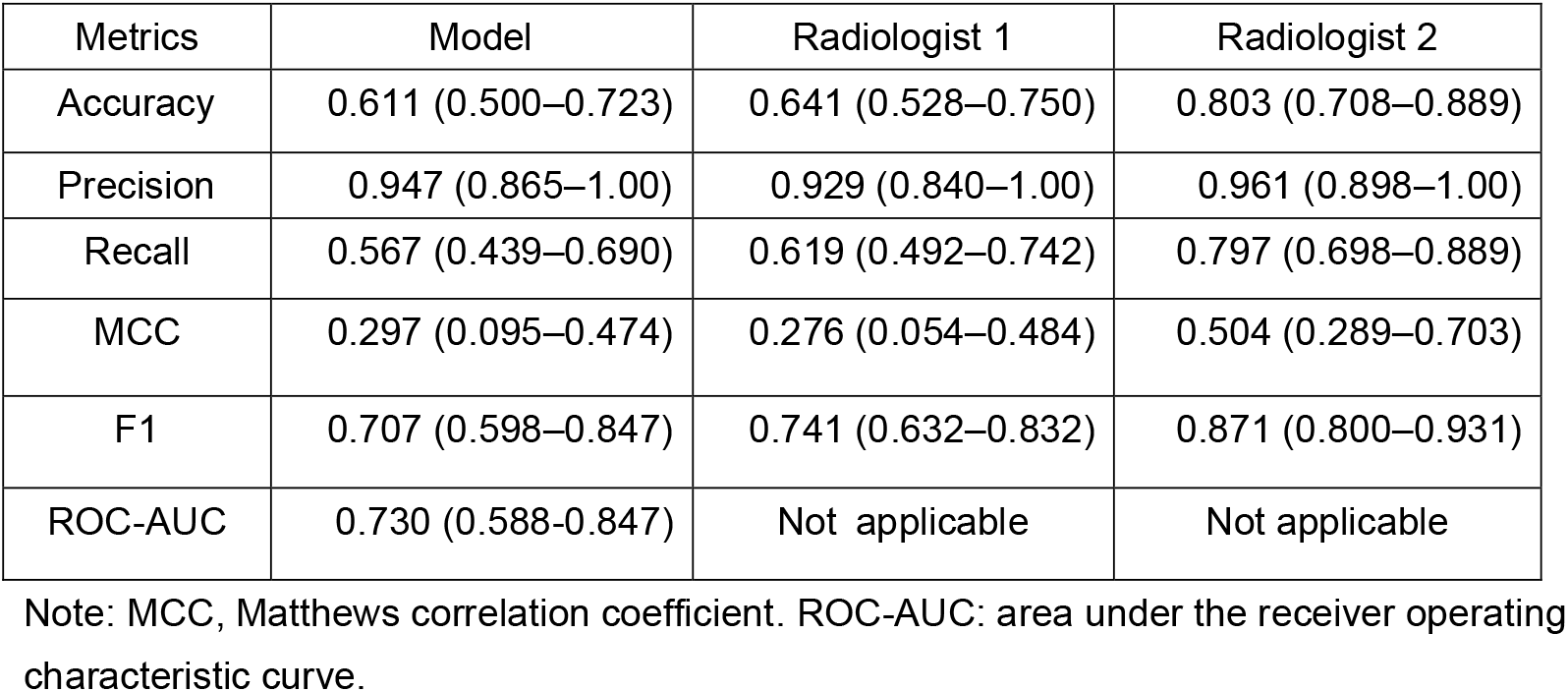
Comparison of Performance Metrics Between the Constructed Model and Two Radiologists (95% Confidence Intervals in Parentheses). ROC-AUC was not applicable for radiologists as they provided binary predictions rather than probability scores.

**Figure 3:**
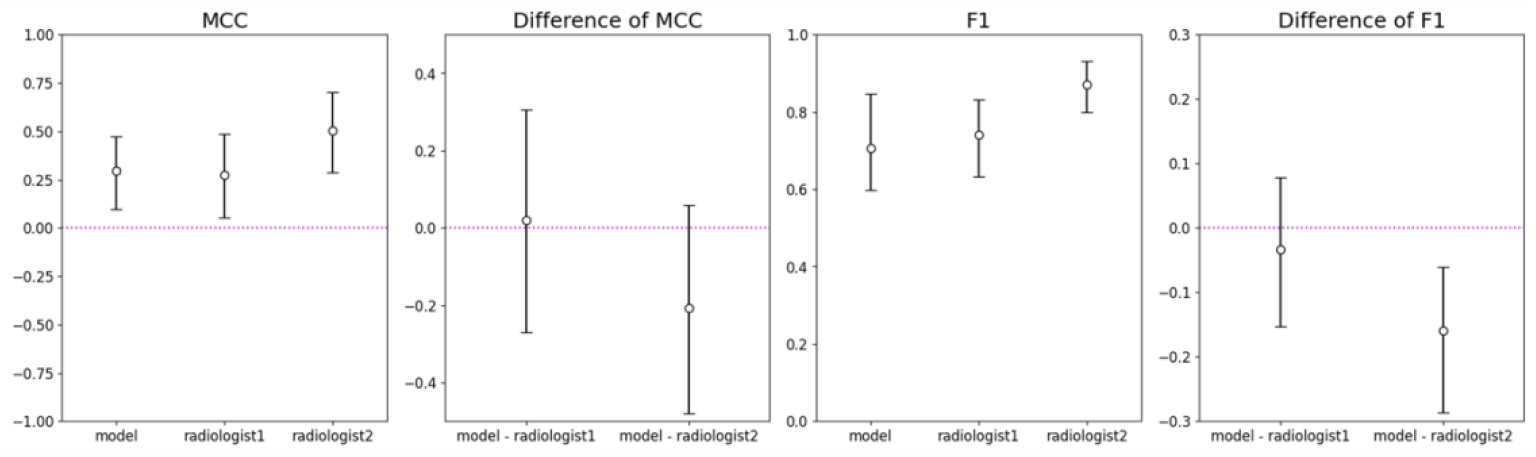
Comparison of MCC and F1 scores between the few-shot learning model and two radiologists for prostate cancer detection, showing absolute values (left panels) and differences between the model and each radiologist (right panels); error bars representing 95% confidence intervals. MCC, Matthews correlation coefficient.

### Visualization of Model Attention

Figure 4 shows the attention visualization of the model using Grad-CAM for prostate cancer classification. The figure shows a 3-channel stacked image, T2WI, and the corresponding GradCAM visualization.

**Figure 4:**
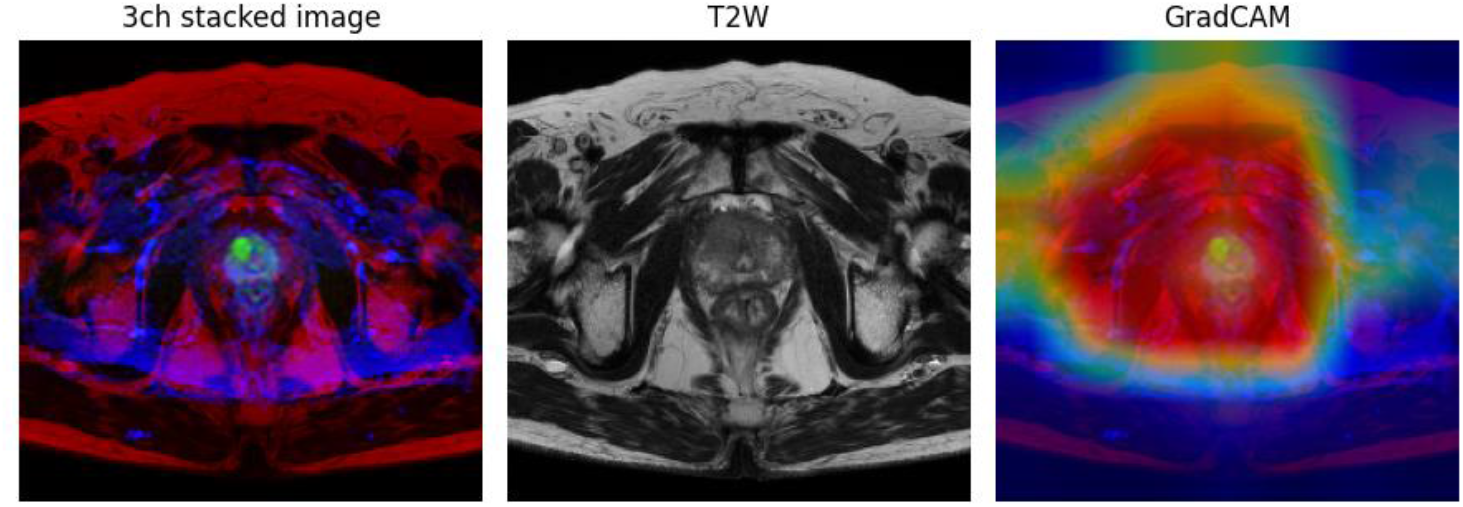
Visualization of prostate MRI analysis, showing (left) the model input as a 3-channel (3ch) stacked image combining T2-weighted images (T2WIs), diffusion-weighted images (DWI), and apparent diffusion coefficient maps (ADC); (center) the original T2WI used for gradient-weighted class activation mapping (GradCAM) overlay; and (right) the resulting GradCAM heatmap highlighting areas of model focus for cancer detection.

The visualization demonstrated that the model focused primarily on the entire prostate gland, as indicated by the warm colors (red and yellow) in GradCAM images. The areas of highest attention aligned with the zonal anatomy of the prostate were visible in the T2-weighted image, suggesting that the model recognizes clinically relevant structures. The surrounding tissues, shown in cooler colors (blue and green), received less attention from the model. This indicates that the model appropriately prioritizes the prostate region in its analysis.

## Discussion

We developed a deep-learning model to classify prostate magnetic resonance images (MRI) based on T2-weighted images (T2WI), diffusion-weighted images (DWI), and apparent diffusion coefficient (ADC) maps to determine whether a biopsy is positive or negative. We devised an algorithm that divides MR images into slice-level sequences and stacks them together, enabling analysis using a two-dimensional model.

Despite training on only 20 cases, the model achieved a classification performance comparable to that of radiologists. The data used in this study consisted of cases in which biopsy was deemed necessary because of remaining suspicion of malignancy based on radiological findings. Many of these cases were difficult to distinguish as benign or malignant; therefore, the results obtained were significantly valuable.

Several deep-learning models have been proposed for the classification of csPCa images. Aldoj and colleagues developed a highly accurate 3D CNN model with an ROC-AUC of 0.897 (23). However, their model required MRI data from a sizable cohort of 175 patients for training purposes. Chen et al. utilized a 2D CNN pretrained on the ImageNet dataset to achieve an AUC of 0.83 (24). They performed training using 330 samples from the PROSTATEx dataset (25). Although it is challenging to directly compare performances because of differences in the target populations, we achieved promising results, with performance comparable to that of a radiologist using only 20 cases.

The main objective of this study was to propose a solution for domain shift (8), which can significantly reduce the accuracy, even for highly accurate models trained on large data sets, because of variations in imaging conditions and imaging equipment. The development of models with sufficient performance, based on a small number of cases at each facility, will enable individual facility-specific model development without considering the domain shift. Furthermore, by utilizing only cases from the facility itself, many issues related to data confidentiality can also be resolved. Here, we developed a high-performance model based on a small number of cases, and established a pathway to resolve these issues.

The future direction of artificial intelligence (AI) in medical imaging is increasingly turning toward the use of foundational models. For instance, BiomedCLIP trained on 15 million text-image pairs from the open PMC dataset has shown remarkable performance in radiology (26). Similarly, CT-CLIP, aimed at becoming a foundational model for lung computed tomography, was developed using scans from more than 20,000 patients and has achieved impressive results (27). The introduction of TotalSegmentator MRI has enabled segmentation of virtually every organ in the body (28). These advancements suggest that the emergence of foundational models or their fine-tuning may lead to the development of AI systems that rely on radiologists’ expertise.

There are only a few reported studies on few-shot learning using MRI. Dhinagar et al. (29) proposed few-shot learning for classifying autism spectrum disorder from MR images. In their approach, they first trained a 3D CNN model using the Autism Brain Imaging Data Exchange dataset (30) and then fine-tuned 20 cases for each site. There are many similarities between their research and ours, such as the use of a small number of data points for fine-tuning (20 cases) and the focus on analyzing the MRI data. We anticipate that performance comparisons between AI and physicians, such as radiologists, under few-shot learning conditions, will become increasingly important. Our research laid the groundwork for this crucial line of inquiry.

Our study had several limitations. First, the performance was evaluated at a single institution, limiting the generalizability of the results. Further evaluation at multiple institutions will allow for a more comprehensive assessment of the generalizability of the model. However, this study focused on demonstrating the feasibility of developing a model to address domain shifts by using a small amount of data from the same institution. Second, we did not validate whether the combination of the model with radiologists would improve diagnostic accuracy in a clinical setting. The proposed algorithm determines whether each slice contains malignant findings, thereby enabling the presentation of noteworthy slices. Therefore, it is expected that the combination of the model and radiologists may improve the diagnostic accuracy. Third, the evaluation was limited to cases in which biopsies were performed and cases without any suspicious findings for malignancy were not included. To construct a model that targets all cases, including cases where biopsies are not required, it is necessary to consider improvements such as setting target labels when biopsies are necessary.

In conclusion, we developed a slice-based MRI model capable of predicting malignancy in biopsies in only 20 cases. The model achieved a performance similar to one radiologist and partially inferior to another, demonstrating its potential for developing effective diagnostic tools with minimal data. This approach may significantly reduce the annotation costs and resource requirements for model development. Future research should explore application to other diseases and extend beyond classification to segmentation and object detection tasks.

## Data Availability

The data used in this study contain personal information of patients and cannot be made publicly available due to ethical and legal restrictions. However, the data may be available for research purposes upon reasonable request and with appropriate procedures. For inquiries about data access, please contact the corresponding author.

## List of abbreviations

ADC: Apparent diffusion coefficient
AI: Artificial intelligence
CI: Confidence intervals
CNN: Convolutional neural network
DWI: Diffusion-weighted images
GradCAM: Gradient-weighted class activation mapping
T2WI: T2-weighted images

## Notes

### Competing Interest Statement

The authors have declared no competing interest.

### Funding Statement

This study did not receive any funding.

### Author Declarations

Ethics Committee of Saitama Medical University International Medical Center gave ethical approval for this work.

